# New Reference Values for Thyroid Volume by Ultrasound in German Children and Adolescents Under Iodine-Sufficient Conditions from a Population-Based Study

**DOI:** 10.1101/2024.01.31.24301932

**Authors:** Raphael Hirtz, Roma Thamm, Ronny Kuhnert, Klaus-Peter Liesenkötter, Michael Thamm, Corinna Grasemann

## Abstract

**Context:** Reliable reference values for thyroid ultrasound measurements are essential to effectively guide individual diagnostics and direct health care measures at the population level, such as iodine fortification programs. However, the latest reference values for total thyroid volume (Tvol) provided by the WHO in 2004 only apply to the 6 to 12-year-old age group and are limited to countries with a long history of iodine sufficiency, which does not reflect the situation in most European countries, including Germany.

**Objective:** The present aims to derive up to date thyroid volume ultrasound reference values in German children and adolescents.

**Design:** Data from the baseline assessment of a nationwide study in German children and adolescents (KiGGS) conducted between 2003 and 2006 were used to determine sex-specific reference values for Tvol in thyroid-healthy participants aged 6 to 17 years by age and body surface area (BSA) according to the Lambda-Mu-Sigma (LMS) method.

**Results:** Data from 5559 participants were available for reference chart construction (girls: 2509 (45.1%)). On average, the 97th percentile is 33.4% and 28.5% higher than the corresponding WHO’s reference values for boys and girls, respectively. These findings are consistent with most other studies in German and European children and adolescents at a similar time of investigation. Notably, the sample used for this study was iodine-sufficient according to WHO criteria.

**Conclusions:** The reference values provided by the WHO are overly conservative for this population and could potentially apply to apply to other European countries with a similar history of iodine supply.

## Introduction

The presence of goiter indicates thyroid disorders of several causes, including iodine deficiency. This may guide further diagnostic procedures at the individual level and help to steer health care measures at the population level, i.e., iodine fortification programs ^1^. Palpation of the thyroid gland for diagnosing goiter suffers from low sensitivity and specificity ^1^. In contrast, thyroid ultrasound is a reliable and replicable method to detect goiter when operated by a trained and experienced examiner ^2,3^. Moreover, the non-invasive nature and short duration make ultrasound studies an ideal diagnostic tool in children and adolescents as long as valid reference values are available. However, thus far, only norms for children and adolescents aged 6 to 12 years have been provided by the WHO ^2^. Moreover, as discussed by the WHO, these reference values may provide a rather conservative estimate of total thyroid volume (Tvol). Additionally, although the reference cohort for the norm values was considered healthy, only urinary iodine concentration from spot samples was determined, without assessing any other thyroid-related parameter. While it has been suggested that spot urinary iodine is a reliable indicator of a population’s iodine supply^4^, it reflects only the iodine intake over the last hours to days. Therefore, it is not suitable for indicating long-term iodine supply at the individual level ^5^. Consequently, there is ambiguous evidence of a relationship between spot urinary iodine concentration (UIC) and thyroid hormone levels ^5^ as well as thyroid volume ^6,7^. In addition, concerning Europe, only Swiss children and adolescents, likely not representative of other European countries, were recruited for the reference sample. In contrast to many European countries that were iodine-deficient until the late 1980s/early 1990s, Switzerland has a long-standing history of iodine sufficiency ^8–10^. However, long-term iodine supply has been suggested to affect Tvol ^2^.

Concerning Tvol reference values for German children and adolescents, there has been a notable absence of a representative study since the reunification of Germany, implying a high risk of bias in Tvol findings from studies confined to specific regions by local confounding ^11^.

Considering these limitations, the present study set out to establish reference values considering important demographic (age, sex) and auxological (body surface area (BSA)) parameters by using data from a large and population-based sample of German children and adolescents aged 6 to 17 years at a time of longer-standing iodine sufficiency ^12^.

## Methods

The German Health Interview and Examination Survey for Children and Adolescents (KiGGS) is a nationally representative study on the health status of German children and adolescents. Details on the study design, the sampling strategy, and the study protocol, including the physical examination of participants, the administered questionnaires and laboratory studies have previously been described elsewhere ^12^ and are detailed in the Supplementary Material 1. ^13^.

The baseline study was conducted by the Robert Koch Institute (RKI) between 2003 and 2006 and included 17,641 participants (7,538 females and 7,485 males | 0-5 years: 5,674, 6-12 years: 7220, 13-17 years: 4,746). The study was approved by the ethics committee of the Charité Berlin (No. 101/2000) and complied with regulations of the Federal Data Protection Act (BDSG). Written informed consent was obtained from families willing to participate. For the present study, a thyroid-healthy sample of KiGGS participants with an ultrasound examination of the thyroid gland was considered. The exclusion criteria, information on missing data, and initial and final sample sizes after exclusion of ineligible participants are detailed in the study flowchart (Figure 1).

**Figure 1.**
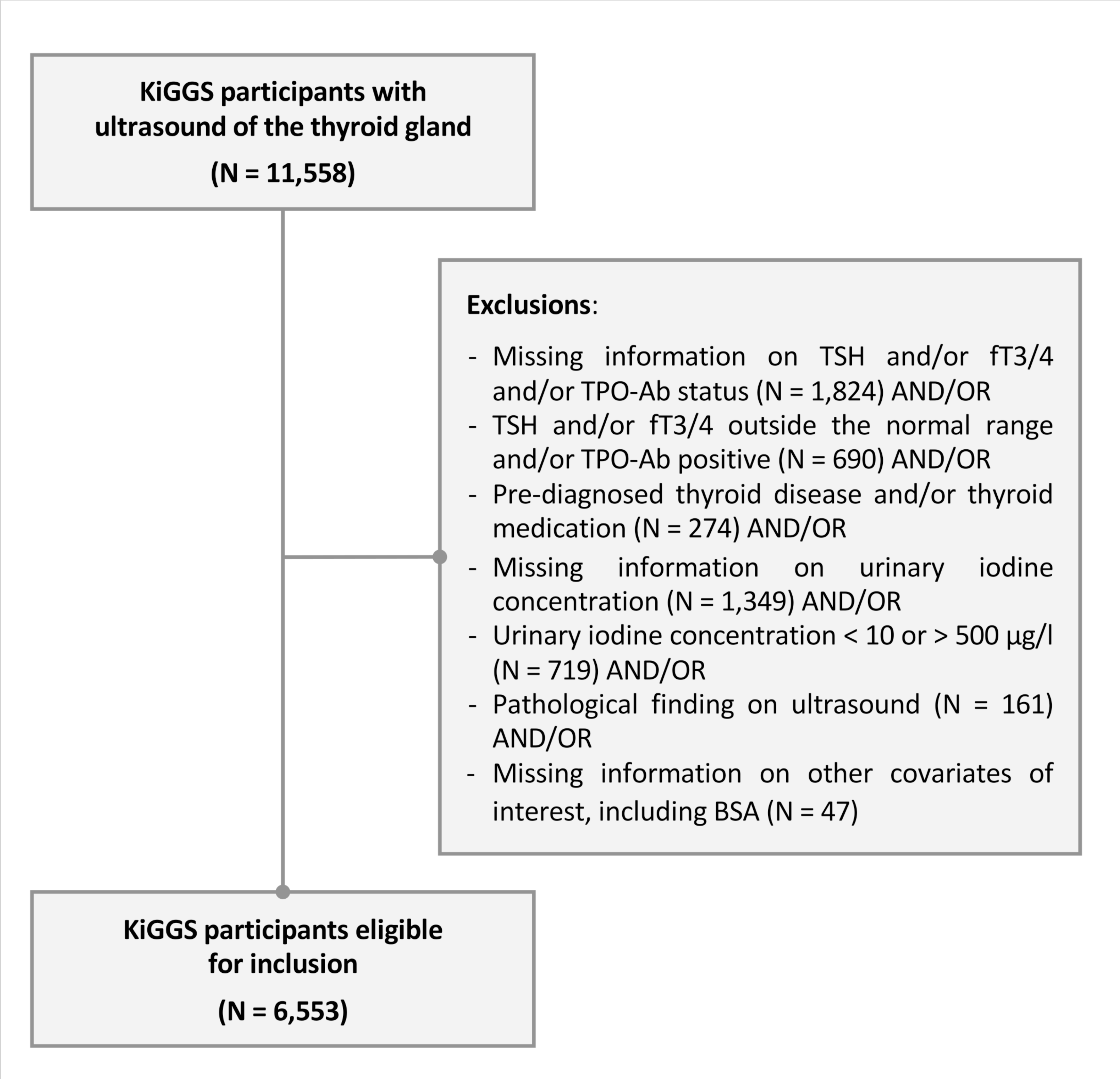
– Study design flowchart. BMI = body mass index | BSA = body surface area | TSH = thyroid-stimulating hormone | fT4 = levothyroxine | fT3 = triiodothyronine. Pathological findings on ultrasound included cysts, nodules, and irregular echogenicity. Note: Some participants had missing information on more than one variable.

### Ultrasound

In children aged 6 years and older, thyroid volume was measured by ultrasound. The length (*l*), width (*w*), and depth (*d*) of each lobe were measured in transverse and longitudinal views by a 7,5 MHz linear-array transducer in supine position. The volume of each lobe was calculated according to Brunn: *l* x *w* x *d* x 0.479. Total thyroid volume was determined by summing the volume of both lobes while excluding the isthmus. Prior to the study, all investigators were trained to perform standardized ultrasonography of the thyroid gland. Throughout the study, average measurements of thyroid volume were compared between investigators to identify and correct systematic errors ^14^.

### Laboratory studies

UIC was determined from spot urine samples according to the Sandell-Kolthoff reaction as recommended by the WHO. Measurements were performed on a Miras Cobas Plus analyzer (Roche Professional Diagnostics, Rotkreuz, Germany; details are given in Thamm, et al. ^14^). UIC determined by this approach was converted according to a formula derived from a re-analysis of samples from the RKI central laboratory by inductively coupled plasma-mass spectrometry in the EUthyroid gold-standard laboratory ^15^. Twenty-four-hour urinary iodine excretion was estimated by correcting UIC by age- and sex-specific 24-hour urinary creatinine excretion reference values, which allows for a reasonable iodine status assessment independent of the individual hydration status (details are provided in Johner, et al. ^16^). Moreover, when accounting for a non-urinary iodine loss of 10%, 24-hour urinary iodine excretion was used to evaluate the percentage of KiGGS participants meeting the age-specific recommended daily allowance (RDA) for iodine, i.e., the average daily intake level required to satisfy the nutrient needs of nearly all (97%–98%) healthy individuals ^17^.

Further information on the assays and their characteristics employed to determine TSH, free levothyroxine (fT4), free triiodothyronine (fT3), and thyroid peroxidase (TPO) antibody levels are provided in the Supplementary Material ^13^.

TSH and fT4 levels were z-transformed according to age- and sex-specific reference ranges using information on the distribution of TSH, fT4, and fT3 from age- and sex-specific percentile charts published for the KiGGS survey participants ^18^.

### Statistics

Data handling and analyses were performed with SPSS 29.0 (Armonk, NY: IBM Corp.) and the SPSS macro PROCESS (version 4.2), R (version 4.2.1), and the R-packages survey (version 4.2-1), GAMLSS (version 5.4-12), and matrixStats (version 0.63.0). To consider the sample design and design-related effects, all analyses were conducted considering sample weights and the cluster structure of the data or normalized weights, as detailed below.

Since the large sample size of the present study rendered minor findings significant, significant results were also interpreted in terms of effect size after conversion to Cohen’s *d* (small 0.2 ≤ *d* < 0.5, medium 0.5 ≤ *d* < 0.8, large *d* ≥ 0.8) and were deemed meaningful only if there was at least a small effect.

Details on testing the assumptions of the employed statistical procedures and the handling of violations are provided in the Supplementary Material ^13^.

### Validation of Ultrasound Results

To identify investigators with deviating measurements despite continuous training, including feedback from interim analyses, each investigator’s mean Tvol of eligible KiGGS participants (Figure 1) was compared to the pooled results of all other investigators by analyses of covariance (ANCOVA). These analyses considered the sample design, age, sex, BSA, and other potential confounders, including z-standardized BMI (z-BMI), TSH, fT3, and fT4 levels and UIC ^6^. This step of analysis was FDR-corrected for multiple comparisons at *q* < .05.

Prior to analysis, data were winsorized by using normalized weights. Winsorization refers to replacing outliers with predefined values. Following recent recommendations ^19^, outliers were designated by values exceeding ± 2.5 times the median absolute difference (MAD) and were replaced by those corresponding to ± 2.5 times the MAD. The data were separately winsorized according to age and BSA.

### Reference Chart Construction

In the final sample of participants examined by eligible investigators as determined by the previous step of analysis, age- and sex-specific percentile reference values for Tvol were derived by the Lambda-Mu-Sigma (LMS) method. The LMS method is a statistical technique for constructing reference curves that can normalize complex data distributions. This was achieved through the Box-Cox Cole Green distribution, a probability distribution that provides additional flexibility, implemented within the GAMLSS R package. The distribution’s parameters L (λ, for skewness of distribution), M (μ, representing the median), and S (σ, denoting the coefficient of variation) were chosen by selecting the subset of hyperparameters that resulted in the lowest Bayesian information criterion (BIC) after conducting a grid search with 5 degrees of freedom (dfs) for each hyperparameter ^20^. To achieve smoothing, cubic splines were utilized. All analyses were separately performed for age and BSA by sex using normalized weights.

### Iodine Supply and Total Thyroid Volume

In the final sample, the effect of iodine supply defined by UIC and estimated 24-hour urinary iodine excretion on Tvol was examined by bivariate correlation and multivariable regression analyses, the latter also considering sex, age, and BSA.

Previous studies have suggested that sex-specific Tvol differences emerge only by peripubertal ages ^2,21,22^. To further examine whether this observation is explained by puberty-related physical changes reflected by an increase in BSA due to the pubertal growth spurt and more adult stature, we performed an ANCOVA on the relationship between Tvol (dependent variable) and sex and age (independent variables), once with and once without BSA. These analyses were considered exploratory, and the results were deemed significant at *p* < .05.

## Results

### Definition of the Study Population

There were 6,553 eligible, thyroid-healthy KiGGS participants, who were iodine-sufficient according to WHO criteria (<20% with UIC <50 µg/l and <50% with UIC <100 µg/l | Table 1). However, only approximately 40% of KiGGS participants met the RDA for iodine, as determined by the estimated 24-hour urinary iodine excretion (sufficiency: 1 to 8 years ≥90 µg/day | 9 to 13 years ≥120 µg/day | 14 to 18 years ≥150 µg/day). Table 1 details this further, showing a decrease in iodine intake by late childhood. Notably, more females than males failed to reach the iodine RDA (*F*(1, 142) = 40.05, *p* < .001).

**Table 1.**
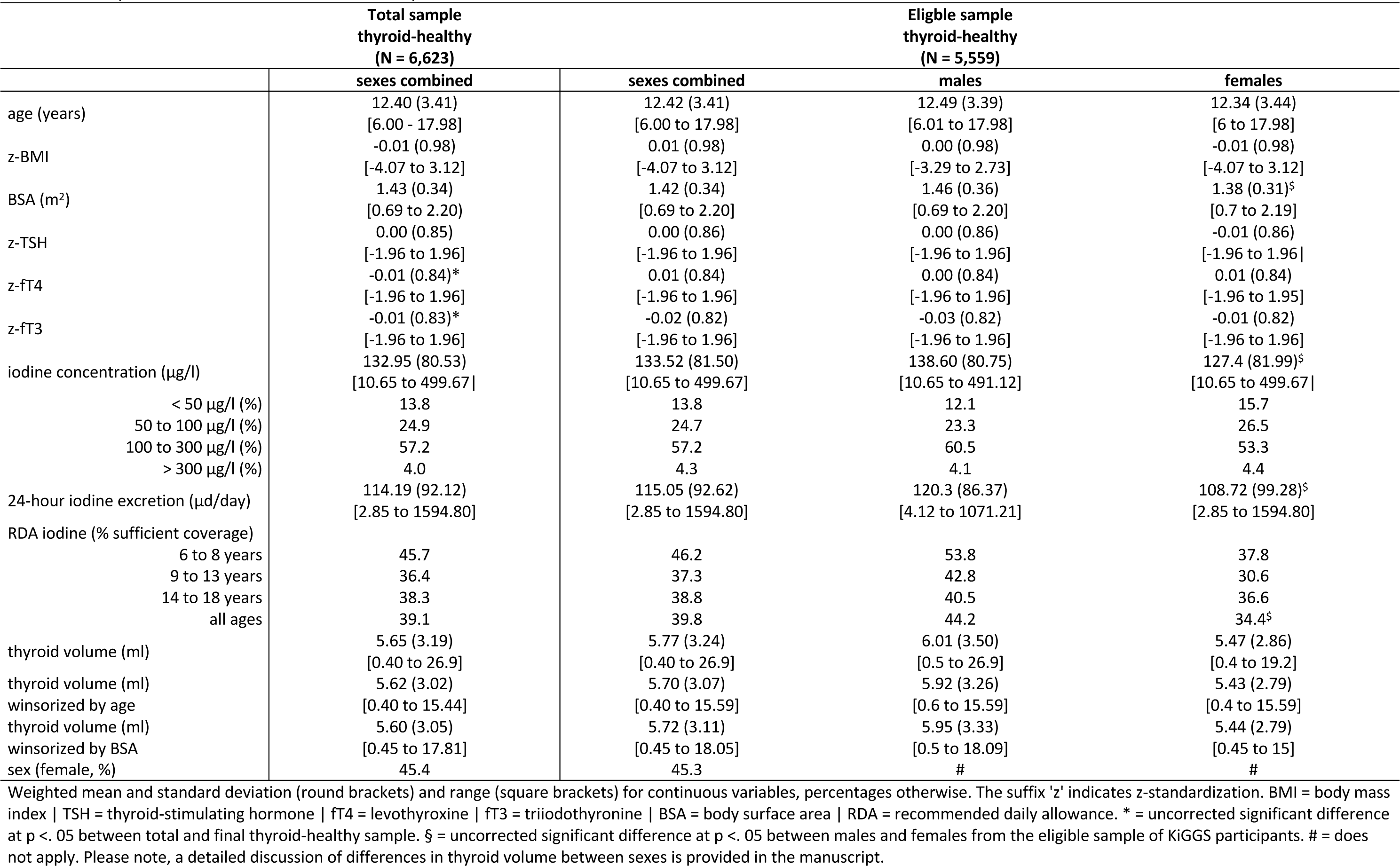
Descriptive Characteristics of the Relevant Samples.

Only 30 participants presented with a BSA above 2.2 m^2^. While these participants were not identified as outliers by their BSA, this number was considered insufficient for reliable reference chart construction beyond 2.2 m^2^. As a consequence, and for consistency with further analyses, these participants were excluded. Thus, Tvol from 231/6,523 (3.5%) and 206/6,523 (3.1%) participants was winsorized by age and BSA, respectively, which is well below a recommended threshold of 5% ^23^.

Despite continuous training and monitoring of the average Tvol of each examiner, there was a significant difference in Tvol adjusted for the study design, sex, age, BSA, z-BMI, z-TSH, z- fT4, z-fT3, and UIC between examiners (Tvol winsorized data by age: *t*(160) = −2.30, *p* = .02 | Tvol winsorized data by BSA: *t*(160) = −2.25, *p* = .03; Table 2). By comparing the mean Tvol of each examiner with the pooled results from all other examiners, examiners 3, 5, 6, and 8 were identified to provide deviant Tvol assessments (Table 2, Supplementary Table 1 ^13^). However, the effect sizes concerning the results of examiner 3 were negligible (Supplementary Table 1 ^13^, *d* = 0.12), indicating that the significance of these findings was driven by the large sample size.

**Table 2.** Thyroid Volume (ml) in the Total Sample of Thyroid-Healthy Participants by.

| Examiner | N | Winsorized by age |  | Winsorized by BSA |  |
| --- | --- | --- | --- | --- | --- |
|  |  | Mean | Mean<sub>adjusted</sub> | Mean | Mean<sub>adjusted</sub> |
| 1 | 892 | 5.73 | 5.72 | 5.73 | 5.72 |
| 2 | 651 | 5.35 | 5.53 | 5.35 | 5.53 |
| 3 | 2,005 | 5.83 | 5.84 | 5.87 | 5.86 |
| 4 | 1,778 | 5.58 | 5.54 | 5.60 | 5.56 |
| 5 | 610 | 4.59 | 4.78 | 4.59 | 4.77 |
| 6 | 192 | 6.79 | 6.46 | 6.80 | 6.47 |
| 7 | 153 | 6.27 | 5.92 | 6.29 | 5.94 |
| 8 | 162 | 4.38 | 4.35 | 4.41 | 4.38 |
| 9 | 80 | 5.85 | 5.67 | 5.86 | 5.68 |

Mean and adjusted mean (considering sex, age, BSA, z-BMI, z-TSH, z-fT4, z-fT3, and iodine excretion) for thyroid volume (ml) by examiner for winsorized data according to age and body surface area (BSA). N = Unweighted number of studied participants by each examiner.

After the exclusion of 964 participants (14.7%) studied by examiners 5, 6, and 8, the final sample for reference chart construction included 5,559 participants. The excluded participants did not differ from this sample regarding any of the characteristics provided in Table 1 when considering a minor effect size concerning z-fT4 (|difference|: 0.01, *t* (160) = 2.09, *p* = .04, d = 0.11) and z-fT3 (|difference|: 0.01, t(160) = −2.58, *p* = .01, d = 0.12 | Table 1). There was no difference between the adjusted mean Tvol by the remaining examiners, including examiner 3, concerning the winsorized data by age (*t*(142)= −0.18, *p* = .86) or BSA (*t*(142)= −0.10, *p* = .92).

### Reference chart construction

Winsorized data (age: 200/5,559 – 3.6%) | BSA: 165/5,559 – 2.9%) from the final sample were used for reference chart construction (dfs for λ, μ, and σ providing the best model fit according to age and BSA by sex are provided in Supplementary Table 2 ^13^). Considering this sample of 5,559 KiGGS participants, Table 3 gives information on the available number of participants for reference chart construction by age in total years (N_median_: 469.5, N_min_: 398, N_max_: 573) and by BSA in increments of 0.2 m^2^ (N: 874, N_min_: 62, N_max_: 1061).

**Table 3.** Sample sizes for categorized age and body surface area.

| **age (years)** | **N** | **BSA (m<sup>2</sup>)** | **N** |
| --- | --- | --- | --- |
| 6 | 402 | 0.6 to 0.8 | 62 |
| 7 | 423 | 0.8 to 1.0 | 796 |
| 8 | 468 | 1.0 to 1.2 | 1,061 |
| 9 | 472 | 1.2 to 1.4 | 952 |
| 10 | 431 | 1.4 to 1.6 | 1,012 |
| 11 | 573 | 1.6 to 1.8 | 1,006 |
| 12 | 522 | 1.8 to 2.0 | 524 |
| 13 | 524 | 2.0 to 2.2 | 146 |
| 14 | 475 |  |  |
| 15 | 471 |  |  |
| 16 | 400 |  |  |
| 17 | 398 |  |  |

As shown in Figure 2A and B and Table 4, there was a continuous increase in Tvol for boys up to the age of 17 years, a pattern also observed in girls, albeit with a slowing trend at advanced ages (for more detailed reference charts concerning age, please refer to Supplementary Table 3 ^13^). Additionally, although girls had larger Tvol during early puberty, this trend reversed later on (Figure 2A and B, Figure 3A). This effect was partially mediated by BSA, as shown in Figure 3A and 3B. Figure 3A shows that a significant sex difference in Tvol begins to manifest by the age of 12. However, when BSA was considered in the relationship between sex and Tvol by an ANCOVA, this effect was lost during mid-puberty (ages 14-16 years) when boys had a larger BSA than girls (Figure 3C), as depicted in Figure 3B.

**Figure 2.**
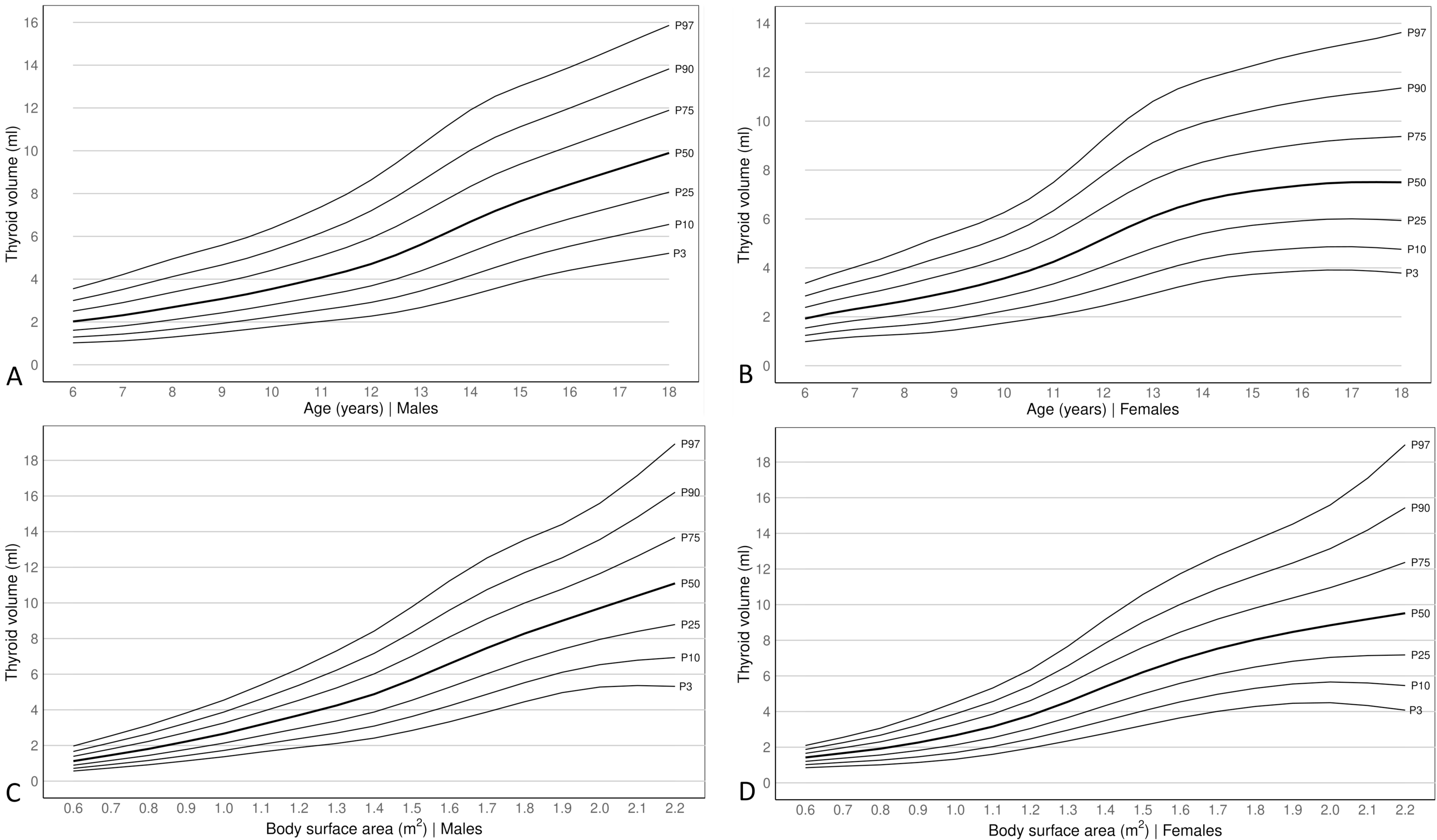
– Total thyroid volume reference curves. References curves with common percentiles (P) for thyroid volume in ml according to age (years) and BSA (m^2^) for males (A/C) and females (B/D).

**Figure 3.**
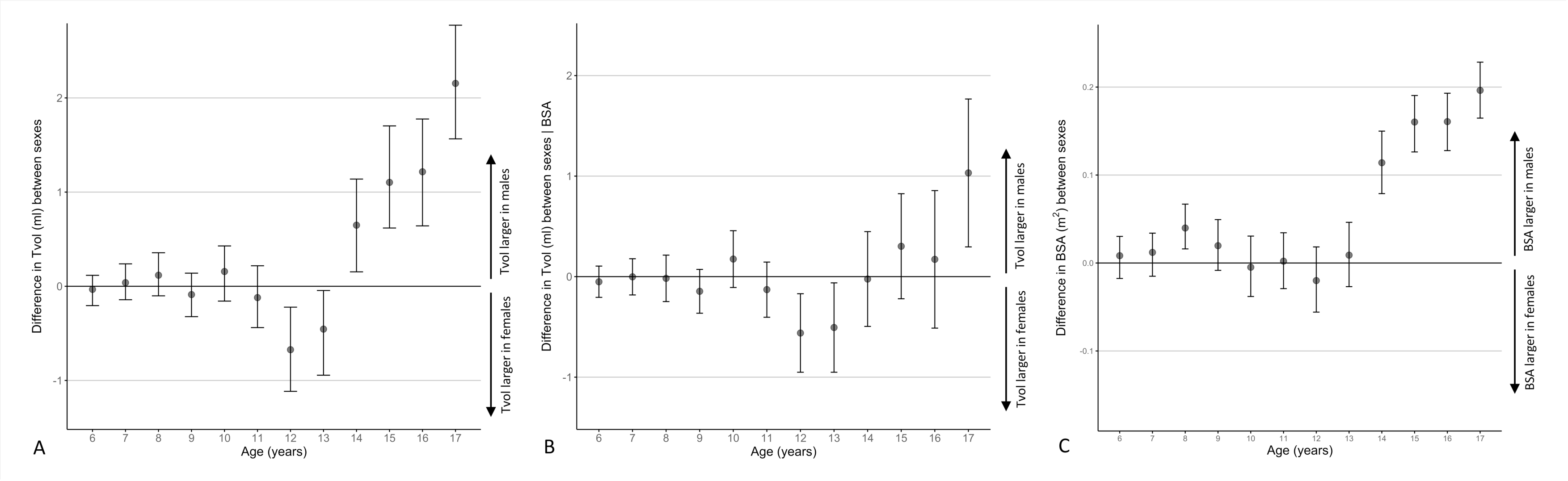
– Total thyroid volume and the mediating effect of BSA. (A) Difference between sexes for total thyroid volume (Tvol) in relation to age (in years) | (B) Difference between sexes for total thyroid volume (Tvol) in relation to age (in years) while considering body surface area (BSA - m^2^) by an analysis of covariance. (C) Difference in BSA between sexes in relation to age. Error bars represent 95% confidence intervals (CI).

**Table 4.**
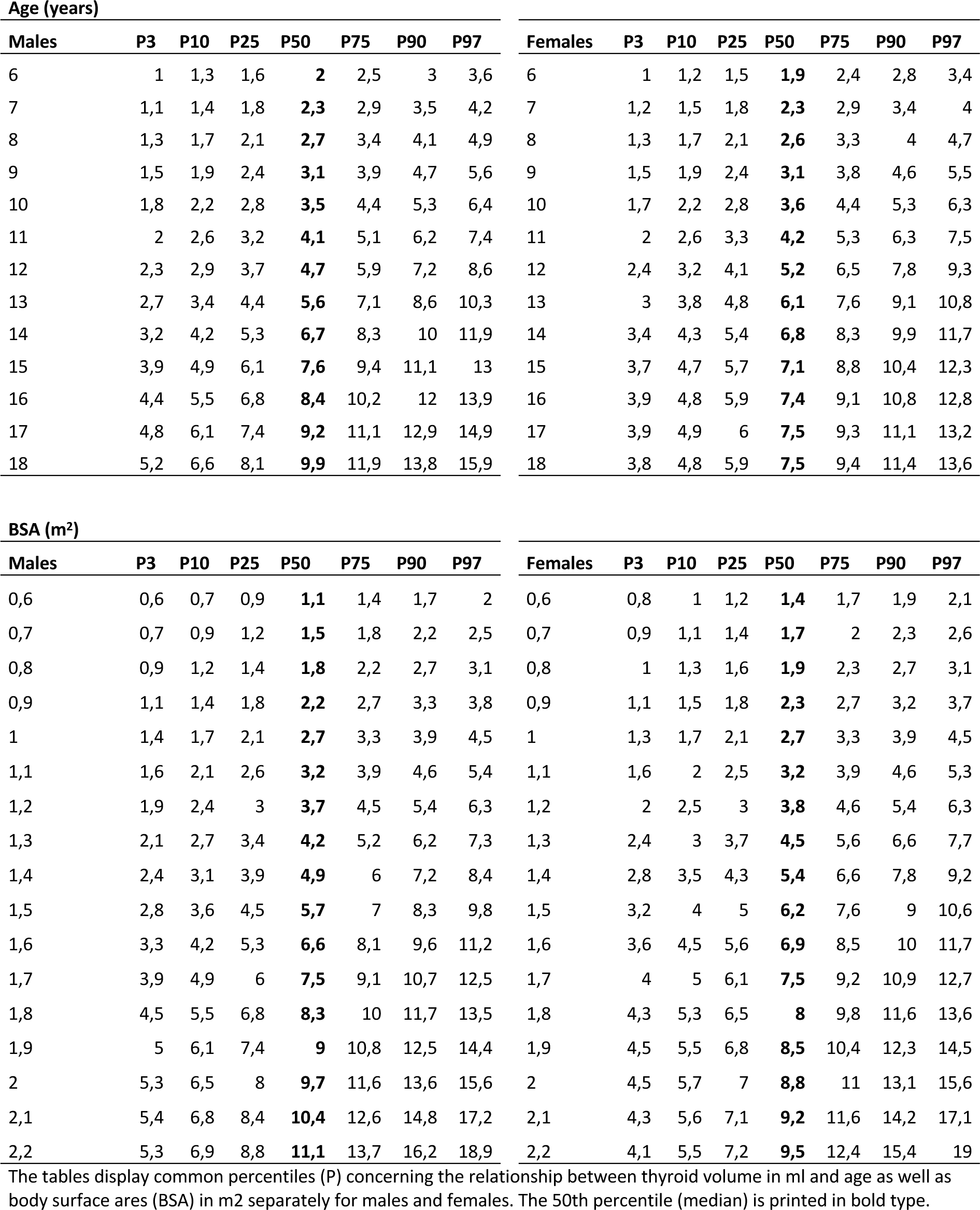
Thyroid Volume (ml) by Age and BSA (m2) Separately in Males and Females.

On average, the 97^th^ percentile is 33.4% and 28.5% higher than the corresponding WHO’s reference values for boys and girls between ages 6 and 12 years in this study, respectively, and a similar picture is seen concerning the 50^th^ percentile (boys: 34.1% - girls: 28.6% | Supplementary Table 4 and Figure 4A^13^). Additionally, concerning BSA, Tvol was higher in the present study than according to the WHO’s reference values (50^th^ percentile – boys: 32.5%, girls: 32.1% | 97^th^ percentile – boys: 17.3%, girls: 25.6% | Figure 4B).

**Figure 4.**
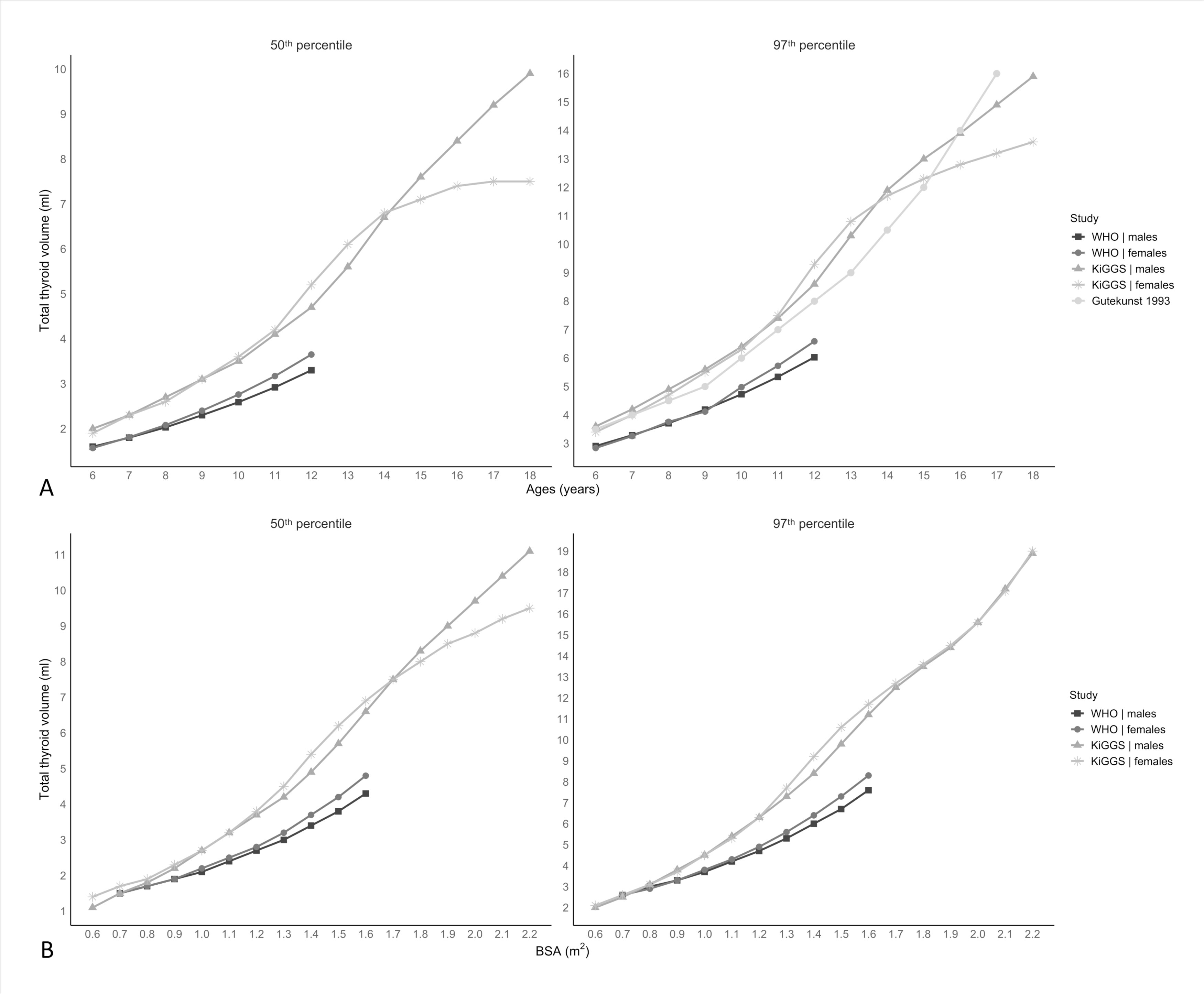
–. Comparison of total thyroid volume between the present and previous studies. (A) Comparison of total thyroid volume (Tvol) according to age (years) in the present study (KiGGS) with the WHO’s reference values and the findings from Gutekunst et al., separately for the 50^th^ and 97^th^ percentile. (B) Comparison of total thyroid volume (Tvol) according to body surface area (BSA – m^2^) in the present study (KiGGS) with the WHO’s reference values, separately for the 50^th^ and 97^th^ percentile.

### Contribution of Iodine Supply to Total Thyroid Volume

It has been suggested that the estimated 24-hour urinary iodine excretion is a better marker of iodine supply than the UIC ^5,16^. This is supported by the bivariate analyses of our study, as there was a closer relationship between the former and thyroid volume (*r* = 0.18, *p* < .001, *d* = 0.37) than with the latter (*r* = 0.05, *p* < .001, d = 0.10), even though both measures of iodine supply were closely related (*r* = 0.58, *p* < .001, *d* = 1.42). However, in contrast to the estimated 24-hour urinary iodine excretion, UIC was found to be (marginally) associated with Tvol in multivariable analyses (Tvol winsorized by age – UIC: b = 7 x 10^-4^, *t*(142) = −1.73, *p* = .09, R^2^ = 3 x 10^-4^; estimated 24-hour iodine excretion: b = 7 x 10^-4^, *t*(142) = −1.46, *p* = .15, R^2^ = 3 x 10^-4^ | Tvol winsorized by BSA – UIC: b = 8 x 10^-4^, *t*(142) = −2.04, *p* = .04, R^2^ = 4 x 10^-4^; estimated 24-hour iodine excretion: b = 7 x 10^-4^, *t*(142) = −1.62, *p* = .11, R^2^ = 5 x 10^-4^ | Supplementary Table 5 ^13^). Nonetheless, neither measure accounted for a significant proportion of variance in Tvol beyond sex, age, and BSA, implying that the (marginal) significant findings concerning UIC were attributable to the large sample size.

## Discussion

The present study provides updated reference charts for Tvol for German children and adolescents at a time of iodine sufficiency based on a large and population-based sample derived from the KiGGS baseline study.

### Time to Shape Up - Comparison to German, European, and International Reference Charts

Liesenkötter, et al. ^11^ studied 1080 children and adolescents aged 3 to 15 years from Berlin in 1997 and reported lower Tvols concerning the 97^th^ percentile in boys over all ages (on average 16%) than in the present study. In contrast, in girls, Tvol was higher during late childhood and lower during pubertal ages than in the present study. However, this study acknowledges local differences in iodine supply and thyroid volume and was intended to provide regional reference values for Tvol.

Moreover, this study presents contradictory findings compared to two other regional studies conducted around the same time in Southern Germany ^24^ with 591 children and adolescents aged 7 to 18 years, and Eastern Germany ^25^ with 2,906 adolescents aged 10 to 18 years. These studies reported higher reference values for the median and the 97th percentile of thyroid volume (Tvol) beginning at pubertal ages (≥ 13 years), unlike the findings of the study by Liesenkötter, et al. ^11^. In particular, the latter study by Hampel, et al. ^25^, a comprehensive state-wide investigation, yielded Tvol values closely mirroring those found in the present study.

As detailed in the introduction, there are several limitations to the current WHO reference charts, including limited coverage beyond 12 years of age and likely a rather conservative estimate of the median and upper limit of Tvol, as discussed by the WHO. For example, these values are lower than those from studies that caused a revision of the WHO’s 1997 reference values by observing significantly smaller Tvol in iodine-sufficient children and adolescents from different countries ^26,27^. In contrast, the findings from these studies are very close, if not identical to the results of the present study, which are approximately 30% higher than the current WHO’s reference values for median Tvol by age and BSA. Moreover, higher reference values than those suggested by the WHO are also implied by findings from studies in other European countries with a history of iodine supply ^15,28^ similar to that of Germany ^21,22^ and even those European countries with long-standing iodine sufficiency ^29^: The present reference values are most closely paralleled by a study at a similar time in 796 Swedish children and adolescents aged 6 to 13 years, a country with iodine fortification as early as 1936 and documented iodine-sufficiency by the late 1980s ^28,29^. Furthermore, the findings of the present study are also consistent with the results of a recent study in 217 Spanish children and adolescents aged 3 to 14 years ^22^. Thus, the reference charts from the present study may serve as a template for other European countries with a longer-standing history of iodine sufficiency when considering its methodological advantages, including the population-based character of the sample, sample size, and information from comprehensive laboratory studies.

### Sex Differences in Total Thyroid Volume

There was a continuous increase in Tvol for boys up to the age of 17 years. However, this pattern is also observed for girls, with a decelerating trend occuring at advanced ages. This finding is reflected by studies in (young) adults, which showed an increase in Tvol up to the age of 30 years in Caucasian ^30^ and up to the age of 50 years in non-Caucasian populations ^31^. Additionally, although girls have a larger Tvol during early pubertal ages, boys have a larger Tvol during advanced ages. We found that BSA partially mediates this observation during peripubertal ages. However, the contribution of sex steroids to these findings beyond their effect on BSA through puberty-related bodily changes remains to be determined. Although testosterone likely increases TSH secretion ^32,33^, explaining the larger Tvol in boys during advanced pubertal stages, the effect of estrogen may differ and vary throughout the lifespan34.

### Practical Implication

Considering that thyroid ultrasound is also used to guide clinical decision making, e.g., on levothyroxine replacement therapy in patients with subclinical hypothyroidism and autoimmune thyroiditis, the present study has implications affecting the clinical management of these patients. Since a slightly enlarged thyroid gland according to previous WHO reference values will now be considered normal, these new reference charts will contribute to a more targeted management by preventing unnecessary replacement therapy when considering that subclinical hypothyroidism spontaneously resolves in approximately ¾ of children and adolescents within few months ^35^ and that mild thyroid autoimmunity with otherwise unremarkable findings, including laboratory studies, likely constitutes a transient phenomenon without clinical implications ^36^.

There was no substantial relationship between spot UIC or estimated 24-hour iodine excretion and Tvol, consistent with the literature on this relationship ^6^ (reasons for this finding are discussed in the Supplemental Material ^13^). Consequently, this finding confirms that spot urinary iodine concentration should not be used in the individual assessment of patients with suspected thyroid pathology.

### Limitations

While it was not possible to quantify the inconsistencies in Tvol measurements among examiners, the methodology utilized in this study to identify examiners with deviating Tvol measurements has been employed in previous research ^37^. In total, three examiners were removed from the study due to diverging Tvol measurements. However, given their random distribution across the country and the large distances between study locations, representativeness of this study remained largely unaffected. This is true for all but one of 16 states and sex (please refer to Supplementary Table 6 ^13^). Importantly, the latter was addressed by sex-specific reference curves. Robustness to the exclusion of examiners is also provided by the absence of any significant difference between the eligible sample and the final sample (used to construct the reference charts) concerning this study’s key characteristics (Table 1).

A recent study revealed a progressive increase in iodine supply in Germany from 1993, peaking around 2003, remaining stable until 2012, and then declining until the latest evaluation in 2018 ^10^. This has been attributed to a reduction in the use of iodized salt by food manufacturers and distributors. Nonetheless, with the identification of this downward trend, remedial strategies have been proposed by the German Federal Institute of Risk Assessment and Federal Ministry of Food and Agriculture ^38^, likely ensuring the validity of the present findings. However, this remains to be determined.

## Data availability

The datasets for this article are not publicly available as the results reported are based on a secondary analysis of data provided by the Robert Koch Institute (RKI), Germany. Requests to access the datasets should therefore be directed to the RKI.

## Author contribution statement

RH conceptualized the present study, analyzed and interpreted the data, and wrote the manuscript. MT and KPL contributed to designing the study and collecting the data. RK provided support concerning data management. All authors participated in scientific discussions and revised the manuscript. All authors approved the submitted version of the manuscript.

