## Supplementary Material for "New Reference Values for Thyroid Volume by Ultrasound in German Children and Adolescents Under Iodine-Sufficient Conditions from a Population-Based Study"

**Overview**

Supplementary Methods 1 – Study Protocol | Physical Examination
Supplementary Methods 2 – Study Protocol | Questionnaires
Supplementary Methods 3 – Study Protocol | Laboratory studies
Supplementary Methods 4 – Study Protocol | Testing of Statistical Assumptions
Supplementary Discussion –
Supplementary Table S1 – Results from the Comparison of Adjusted Mean Thyroid Volume between Examiners
Supplementary Table S2 – Degrees of freedom for optimal LMS model fit
Supplementary Table S3 – Detailed Reference Charts for Total Thyroid Volume by Age
Supplementary Table S4 - Comparison of Total Thyroid Volume - KiGGS vs. WHO
Supplementary Table S5 – ANCOVA - Total Thyroid Volume and Iodine Supply
Supplementary References
Supplementary Table S6A - Comparison Between Total and Final Sample Concerning Important Characteristics Determining Representativeness
Supplementary Table S6B - Comparison Between Total and Final Sample Concerning Important Characteristics Determining Representativeness Stratified by Sex

### Supplementary Methods

#### Study Protocol

#### Physical examination

In children 2 years and older, height was determined in upright posture using a calibrated stadiometer without wearing shoes. Height was recorded with a precision of 0.1 mm. Weight was measured wearing underwear by an electronic scale displaying weight with a precision of 0.1 kg [1]. BMI was determined by the ratio of weight in kg and the height in meters squared ($kg/m^2$) and was z-standardized according to percentile charts for German children and adolescents [2]. BSA was computed according to: weight $(kg)^{0.425}$ * height $(cm)^{0.725}$ * 71.84 *$10^{-4}$.

#### Questionnaires

Participants aged 11 years and older and parents of minor participants completed self-administered, standardized questionnaires. Moreover, parents of minors took part in a computer-assisted personal interview (CAPI) conducted by a specially trained study physician. The CAPI comprised a detailed section on the use of medication within the last 7 days, either prescribed or sold as over-the-counter drug. To verify the reported medication, parents were asked to bring the original containers or package inserts on the day of the interview. Specific ATC (Anatomical Therapeutic Chemical) codes were recorded for all reported medication. The interview also covered the participants' medical history and asked for physician-diagnosed diseases and chronic conditions [3].

#### Laboratory studies

Blood samples were obtained by venous puncture after a median fasting period of 2 hours using a vacutainer system. Whole blood was stored at 4°C and serum at – 40°C before sample transfer to the central laboratory of the RKI within 3 days after sample collection [4].

Analyses of serum TSH and fT4 levels were performed with the Elecsys 2010® immunoassay analyzer (Roche Professional Diagnostics, Rotkreuz, Germany) and sandwich electrochemiluminescence immunoassays (ECLIA) with an inter-assay variation of <3.9% and <5.3% and a detection range of 0.005 to 100 IU/ml and 0.023 to 7.77 ng/ml, respectively [4]. TPO-antibody levels were measured on a Phadia 250® immunoassay system (Thermo Fisher Scientific, Uppsala, Sweden) in conjunction with the ImmunoCap TPO assay, a fluoro-enzymeimmunoassay (FEIA) with an inter-assay variation of <3.9% (at 494 IU/ml). The measurement range of the assay lies between 33.4 and 3,600 IU/ml and the cut-off level for TPO-positivity has been established at > 100 IU/ml.

**Testing of Statistical Assumptions**

Normality of residuals was assessed by visual inspection of QQ-plots, heterogeneity of variances across groups by Levene's test (according to the median – ANCOVA analysis), homoscedasticity by plotting predicted values against residuals (regression analysis), and the homogeneity of regression slopes by assessing the interaction between the independent grouping variable and covariates (ANCOVA). Linearity was assessed by bivariate scatterplots. When assumptions were violated, analyses were performed by bootstrapping with 1,000 bootstrap samples.

### Supplementary Discussion

#### Contribution of Iodine to Total Thyroid Volume

There was no substantial relationship between spot UIC or estimated 24-hour iodine excretion and Tvol, likely for several reasons. Since approximately 90% of iodine is excreted within 24 to 48 hours after ingestion, spot UIC only reflects the iodine intake over the last hours to days [5]. Thus, spot UIC is not an indicator of long-term iodine sufficiency at the individual level due to considerable intraindividual day-by-day variation. Moreover, the WHO cut-off for spot UIC to indicate sufficient iodine supply has been criticized [6], which also applies to the method itself. Spot UICs depend on individual hydration status, and without accounting for daily urinary output, this may result in misleading conclusions [7]. Likely for the same reason, we found iodine sufficiency by spot UIC according to the WHO criteria, while only approximately 40% of the KiGGS participants met the RDA for iodine using the estimated 24-hour iodine excretion.

**Supplementary Table 1** - Results from the Comparison of Adjusted Mean Thyroid Volume between Examiners.

| | Winsorized by age | | | | Winsorized by BSA | | | |
|---|---|---|---|---|---|---|---|---|
| **Examiner** | **b** | **t** | **p** | **d** | **b** | **t** | **p** | **d** |
| 1 | 0.16 | 1.07 | 0.29 | # | 0.14 | 0.93 | 0.35 | # |
| 2 | -0.06 | -0.34 | 0.74 | # | -0.08 | -0.43 | 0.67 | # |
| 3 | 0.36 | 3.22 | 0.002 | 0.12 | 0.37 | 3.26 | 0.001 | 0.12 |
| 4 | -0.07 | -0.54 | 0.59 | # | -0.06 | -0.43 | 0.67 | # |
| 5 | -0.90 | -7.15 | <.001 | 0.33 | -0.91 | -7.23 | <.001 | 0.34 |
| 6 | 0.90 | 4.14 | <.001 | 0.29 | 0.89 | 4.03 | <.001 | 0.29 |
| 7 | 0.35 | 1.06 | 0.29 | # | 0.35 | 1.05 | 0.29 | # |
| 8 | -1.26 | -3.15 | 0.002 | 0.43 | -1.24 | -3.02 | 0.003 | 0.41 |
| 9 | 0.09 | 0.48 | 0.63 | # | 0.09 | 0.34 | 0.73 | # |

Results from the comparison of adjusted (sex, age, BSA, z-BMI, z-TSH, z-fT4, z-fT3, and iodine excretion) mean thyroid volumes between each examiner and the pooled results from all other examiners for winsorized data according to age and body surface area (BSA). b = unstandardized regression coefficient. t = t-value. p = p-value, d = Cohen's d for significant findings. # = does not apply.

**Supplementary Table 2** - Degrees of freedom for optimal model fit (LMS method).

| | **μ (df)** | **σ (df)** | **λ (df)** |
|---|---|---|---|
| age | 5 | 5 | 0 |
| age \| males | 5 | 5 | 1 |
| age \| females | 5 | 5 | 1 |
| BSA | 4 | 5 | 0 |
| BSA \| males | 5 | 5 | 0 |
| BSA \| females | 5 | 5 | 0 |

Degrees of freedom (df) for Box-Cox Cole Green transformation parameters with respect to age and body surface are (BSA), separately for both sexes, providing the best model fit (lowest AIC) for thyroid volume.

**Supplementary Table 3**- Thyroid Volume by Age in Both Sexes and Separately in Males and Females

**Age | Males**

| **age (years)** | **P3** | **P10** | **P25** | **P50** | **P75** | **P90** | **P97** |
|---|---|---|---|---|---|---|---|
| 6 | 1 | 1.3 | 1.6 | **2** | 2.5 | 3 | 3.6 |
| 6.5 | 1.1 | 1.4 | 1.7 | **2.2** | 2.7 | 3.3 | 3.9 |
| 7 | 1.1 | 1.4 | 1.8 | **2.3** | 2.9 | 3.5 | 4.2 |
| 7.5 | 1.2 | 1.5 | 1.9 | **2.5** | 3.1 | 3.8 | 4.6 |
| 8 | 1.3 | 1.7 | 2.1 | **2.7** | 3.4 | 4.1 | 4.9 |
| 8.5 | 1.4 | 1.8 | 2.3 | **2.9** | 3.6 | 4.4 | 5.3 |
| 9 | 1.5 | 1.9 | 2.4 | **3.1** | 3.9 | 4.7 | 5.6 |
| 9.5 | 1.6 | 2.1 | 2.6 | **3.3** | 4.1 | 5 | 5.9 |
| 10 | 1.8 | 2.2 | 2.8 | **3.5** | 4.4 | 5.3 | 6.4 |
| 10.5 | 1.9 | 2.4 | 3 | **3.8** | 4.7 | 5.7 | 6.9 |
| 11 | 2 | 2.6 | 3.2 | **4.1** | 5.1 | 6.2 | 7.4 |
| 11.5 | 2.1 | 2.7 | 3.4 | **4.4** | 5.5 | 6.6 | 8 |
| 12 | 2.3 | 2.9 | 3.7 | **4.7** | 5.9 | 7.2 | 8.6 |
| 12.5 | 2.4 | 3.2 | 4 | **5.1** | 6.5 | 7.8 | 9.4 |
| 13 | 2.7 | 3.4 | 4.4 | **5.6** | 7.1 | 8.6 | 10.3 |
| 13.5 | 2.9 | 3.8 | 4.8 | **6.1** | 7.7 | 9.3 | 11.1 |
| 14 | 3.2 | 4.2 | 5.3 | **6.7** | 8.3 | 10 | 11.9 |
| 14.5 | 3.6 | 4.5 | 5.7 | **7.2** | 8.9 | 10.6 | 12.5 |
| 15 | 3.9 | 4.9 | 6.1 | **7.6** | 9.4 | 11.1 | 13 |
| 15.5 | 4.2 | 5.2 | 6.5 | **8** | 9.8 | 11.6 | 13.5 |
| 16 | 4.4 | 5.5 | 6.8 | **8.4** | 10.2 | 12 | 13.9 |
| 16.5 | 4.6 | 5.8 | 7.1 | **8.8** | 10.6 | 12.4 | 14.4 |
| 17 | 4.8 | 6.1 | 7.4 | **9.2** | 11.1 | 12.9 | 14.9 |
| 17.5 | 5 | 6.3 | 7.8 | **9.5** | 11.5 | 13.4 | 15.4 |
| 18 | 5.2 | 6.6 | 8.1 | **9.9** | 11.9 | 13.8 | 15.9 |

**Age | Females**

| **age (years)** | **P3** | **P10** | **P25** | **P50** | **P75** | **P90** | **P97** |
|---|---|---|---|---|---|---|---|
| 6 | 1 | 1.2 | 1.5 | **1.9** | 2.4 | 2.8 | 3.4 |
| 6.5 | 1.1 | 1.4 | 1.7 | **2.1** | 2.6 | 3.1 | 3.7 |
| 7 | 1.2 | 1.5 | 1.8 | **2.3** | 2.9 | 3.4 | 4 |
| 7.5 | 1.2 | 1.6 | 2 | **2.5** | 3.1 | 3.7 | 4.3 |
| 8 | 1.3 | 1.7 | 2.1 | **2.6** | 3.3 | 4 | 4.7 |
| 8.5 | 1.4 | 1.8 | 2.2 | **2.8** | 3.6 | 4.3 | 5.1 |
| 9 | 1.5 | 1.9 | 2.4 | **3.1** | 3.8 | 4.6 | 5.5 |
| 9.5 | 1.6 | 2.1 | 2.6 | **3.3** | 4.1 | 4.9 | 5.8 |
| 10 | 1.7 | 2.2 | 2.8 | **3.6** | 4.4 | 5.3 | 6.3 |
| 10.5 | 1.9 | 2.4 | 3.1 | **3.9** | 4.8 | 5.8 | 6.8 |
| 11 | 2 | 2.6 | 3.3 | **4.2** | 5.3 | 6.3 | 7.5 |
| 11.5 | 2.2 | 2.9 | 3.7 | **4.7** | 5.9 | 7 | 8.3 |
| 12 | 2.4 | 3.2 | 4.1 | **5.2** | 6.5 | 7.8 | 9.3 |
| 12.5 | 2.7 | 3.5 | 4.4 | **5.7** | 7.1 | 8.5 | 10.1 |
| 13 | 3 | 3.8 | 4.8 | **6.1** | 7.6 | 9.1 | 10.8 |
| 13.5 | 3.2 | 4.1 | 5.1 | **6.5** | 8 | 9.6 | 11.3 |
| 14 | 3.4 | 4.3 | 5.4 | **6.8** | 8.3 | 9.9 | 11.7 |
| 14.5 | 3.6 | 4.5 | 5.6 | **7** | 8.6 | 10.2 | 12 |
| 15 | 3.7 | 4.7 | 5.7 | **7.1** | 8.8 | 10.4 | 12.3 |
| 15.5 | 3.8 | 4.7 | 5.8 | **7.3** | 8.9 | 10.6 | 12.5 |
| 16 | 3.9 | 4.8 | 5.9 | **7.4** | 9.1 | 10.8 | 12.8 |
| 16.5 | 3.9 | 4.9 | 6 | **7.5** | 9.2 | 11 | 13 |
| 17 | 3.9 | 4.9 | 6 | **7.5** | 9.3 | 11.1 | 13.2 |
| 17.5 | 3.9 | 4.8 | 6 | **7.5** | 9.3 | 11.2 | 13.4 |
| 18 | 3.8 | 4.8 | 5.9 | **7.5** | 9.4 | 11.4 | 13.6 |

The tables display common percentiles (P) concerning the relationship between thyroid volume in ml and age in years separately for males and females. The 50th percentile (median) is printed in bold type.

**Table 4A** - Comparison of Total Thyroid Volume in KiGGS with the WHO´s Reference Values (2004) by Age.

**50th percentile - Males**

| Age (years) | 6 | 7 | 8 | 9 | 10 | 11 | 12 |
|---|---|---|---|---|---|---|---|
| Tvol (ml) - KiGGS | 2.0 | 2.3 | 2.7 | 3.1 | 3.5 | 4.1 | 4.7 |
| Tvol (ml) - WHO | 1.6 | 1.8 | 2.0 | 2.3 | 2.6 | 2.9 | 3.3 |
| % Tvol KiGGS with reference WHO | 125.0 | 127.8 | 133.0 | 134.8 | 134.6 | 141.4 | 142.4 |

**97th percentile - Males**

| Age (years) | 6 | 7 | 8 | 9 | 10 | 11 | 12 |
|---|---|---|---|---|---|---|---|
| Tvol (ml) - KiGGS | 3.6 | 4.2 | 4.9 | 5.6 | 6.4 | 7.4 | 8.6 |
| Tvol (ml) - WHO | 2.9 | 3.3 | 3.7 | 4.2 | 4.7 | 5.3 | 6.0 |
| % Tvol KiGGS with reference WHO | 123.7 | 127.7 | 132.1 | 133.7 | 135.3 | 138.6 | 142.6 |

Tvol = Total thyroid volume (in ml). Please note, Tvol is rounded to the first decimal place.

**50th percentile - Females**

| Age (years) | 6 | 7 | 8 | 9 | 10 | 11 | 12 |
|---|---|---|---|---|---|---|---|
| Tvol (ml) - KiGGS | 1.9 | 2.3 | 2.6 | 3.1 | 3.6 | 4.2 | 5.2 |
| Tvol (ml) - WHO | 1.6 | 1.8 | 2.1 | 2.4 | 2.8 | 3.2 | 3.7 |
| % Tvol KiGGS with reference WHO | 118.8 | 127.8 | 123.8 | 129.2 | 128.6 | 131.3 | 140.5 |

**97th percentile - Females**

| Age (years) | 6 | 7 | 8 | 9 | 10 | 11 | 12 |
|---|---|---|---|---|---|---|---|
| Tvol (ml) - KiGGS | 3.4 | 4.0 | 4.7 | 5.5 | 6.3 | 7.5 | 9.3 |
| Tvol (ml) - WHO | 2.8 | 3.3 | 3.8 | 4.1 | 5.0 | 5.7 | 6.6 |
| % Tvol KiGGS with reference WHO | 119.7 | 122.7 | 125.0 | 133.5 | 126.5 | 130.9 | 141.1 |

Tvol = Total thyroid volume (in ml). Please note, Tvol is rounded to the first decimal place.

**Table 4B** - Comparison of Total Thyroid Volume in KiGGS with the WHO´s Reference Values (2004) by Body Surface Area

**50th percentile - Males**

| BSA ($m^2$) | 0.7 | 0.8 | 0.9 | 1.0 | 1.1 | 1.2 | 1.3 | 1.4 | 1.5 | 1.6 |
|---|---|---|---|---|---|---|---|---|---|---|
| **Tvol (ml) - KiGGS** | 1.5 | 1.8 | 2.2 | 2.7 | 3.2 | 3.7 | 4.2 | 4.9 | 5.7 | 6.6 |
| **Tvol (ml) - WHO** | 1.5 | 1.7 | 1.9 | 2.1 | 2.4 | 2.7 | 3.0 | 3.4 | 3.8 | 4.3 |
| **% Tvol KiGGS with reference WHO** | 102.0 | 108.4 | 118.3 | 128.6 | 135.6 | 139.6 | 140.5 | 145.8 | 150.8 | 155.3 |

**97th percentile - Males**

| BSA ($m^2$) | 0.7 | 0.8 | 0.9 | 1.0 | 1.1 | 1.2 | 1.3 | 1.4 | 1.5 | 1.6 |
|---|---|---|---|---|---|---|---|---|---|---|
| **Tvol (ml) - KiGGS** | 2.5 | 3.1 | 3.8 | 4.5 | 5.4 | 6.3 | 7.3 | 8.4 | 9.8 | 11.2 |
| **Tvol (ml) - WHO** | 2.6 | 3.0 | 3.3 | 3.7 | 4.2 | 4.7 | 5.3 | 6.0 | 6.7 | 7.6 |
| **% Tvol KiGGS with reference WHO** | 104.8 | 105.1 | 114.5 | 120.6 | 128.6 | 133.2 | 137.2 | 140.5 | 145.6 | 148.0 |

Tvol = Total thyroid volume (in ml). Please note, Tvol is rounded to the first decimal place.

**50th percentile - Females**

| BSA ($m^2$) | 0.7 | 0.8 | 0.9 | 1.0 | 1.1 | 1.2 | 1.3 | 1.4 | 1.5 | 1.6 |
|---|---|---|---|---|---|---|---|---|---|---|
| **Tvol (ml) - KiGGS** | 1.7 | 1.9 | 2.3 | 2.7 | 3.2 | 3.8 | 4.5 | 5.4 | 6.2 | 6.9 |
| **Tvol (ml) - WHO** | 1.5 | 1.7 | 1.9 | 2.2 | 2.5 | 2.8 | 3.2 | 3.7 | 4.2 | 4.8 |
| **% Tvol KiGGS with reference WHO** | 116.4 | 113.8 | 121.1 | 124.4 | 129.6 | 134.8 | 140.2 | 147.5 | 148.7 | 145.0 |

**97th percentile - Females**

| BSA ($m^2$) | 0.7 | 0.8 | 0.9 | 1.0 | 1.1 | 1.2 | 1.3 | 1.4 | 1.5 | 1.6 |
|---|---|---|---|---|---|---|---|---|---|---|
| **Tvol (ml) - KiGGS** | 2.6 | 3.1 | 3.7 | 4.5 | 5.3 | 6.3 | 7.7 | 9.2 | 10.6 | 11.7 |
| **Tvol (ml) - WHO** | 2.6 | 2.9 | 3.3 | 3.8 | 4.3 | 4.9 | 5.6 | 6.4 | 7.3 | 8.3 |
| **% Tvol KiGGS with reference WHO** | 101.6 | 106.5 | 111.4 | 118.7 | 122.7 | 128.0 | 137.3 | 143.8 | 145.4 | 140,6 |

Tvol = Total thyroid volume (in ml). Please note, Tvol is rounded to the first decimal place.

**Supplementary Table 5** - Total Thyroid Volume and Iodine Supply

| | IV | b | SE | t | p |
|---|---|---|---|---|---|
| **Tvol winsorized by age** | **UIC (µg/l)** | -7 x $10^{-4}$ | 4 x $10^{-4}$ | -1.73 | .09 |
| | sex | -0.05 | 0.06 | -0.75 | .45 |
| | age (years) | 0.21 | 0.02 | 8.74 | <.001 |
| | BSA (m2) | 5.27 | 0.25 | 21.44 | <.001 |
| | **Estimated 24-hour iodine excretion (µg/d)** | -7 x $10^{-4}$ | 5 x $10^{-4}$ | -1.46 | .15 |
| | sex | -0.05 | 0.06 | -0.68 | .50 |
| | age (years) | 0.21 | 0.02 | 8.63 | <.001 |
| | BSA (m2) | 5.33 | 0.26 | 20.61 | <.001 |
| **Tvol winsorized by BSA** | **UIC (µg/l)** | -8 x $10^{-4}$ | 5 x $10^{-4}$ | -2.04 | .04 |
| | sex | -0.05 | 0.07 | -0.76 | .45 |
| | age (years) | 0.17 | 0.03 | 6.19 | <.001 |
| | BSA (m2) | 5.73 | 0.31 | 19.46 | <.001 |
| | **Estimated 24-hour iodine excretion (µg/d)** | -7 x $10^{-4}$ | 5 x $10^{-4}$ | -1.62 | .11 |
| | sex | -0.04 | 0.06 | -0.68 | .50 |
| | age (years) | 0.16 | 0.03 | 8..63 | <.001 |
| | BSA (m2) | 5.33 | 0.31 | 20.61 | <.001 |

The table displays the parameter estimate b, the standard error (SE), the t-value, and the p-value for the models concerning thyroid volume winsorized by age (dependent variable) for the independent variables (IV) outlined in the respective column. The suffix 'z' indicates z-standardization. BMI = body mass index | BSA = body surface area | TSH = thyroid-stimulating hormone | fT4 = levothyroxine | fT3 = triiodothyronine.

**Supplementary Table 6 A**

Comparison Between Total and Final Sample Concerning Important Characteristics Determining Representativeness

| variable | label | $N_{total\ sample}$ = 11,024 | Proportion (%) | $N_{final\ sample}$ = 5,559 | Proportion (%) |
|---|---|---|---|---|---|
| sex | male | 5,666 | 51.32 | 3,050 | 54.78 |
| sex | female | 5,358 | 48.68 | 2,509 | 45.22 |
| age group | 3-6 years | 861 | 7.64 | 402 | 7.17 |
| age group | 7-10 years | 3,739 | 30.70 | 1,794 | 29.36 |
| age group | 11-13 years | 2,894 | 24.44 | 1,619 | 26.27 |
| age group | 14-17 years | 3,530 | 37.22 | 1,744 | 37.21 |
| educational level | basic education | 1,880 | 34.15 | 923 | 33.46 |
| educational level | intermediate education | 5,974 | 44.25 | 3,066 | 45.43 |
| educational level | higher education | 2,959 | 19.96 | 1,472 | 19.64 |
| educational level | NA | 211 | 1.64 | 98 | 1.48 |
| state | Schleswig-Holstein | 324 | 3.56 | 191 | 4.27 |
| state | Hamburg | 136 | 1.76 | 78 | 2.00 |
| state | Lower Saxony | 860 | 10.56 | 496 | 12.10 |
| state | Bremen | 64 | 0.71 | 25 | 0.57 |
| state | Northrhine-Westphalia | 2,079 | 23.14 | 1,097 | 24.50 |
| state | Hesse | 567 | 7.36 | 331 | 8.12 |
| state | Rhineland Palatinate | 447 | 5.14 | 165 | 3.84 |
| state | Baden-Württemberg | 1,285 | 13.90 | 504 | 10.63 |
| state | Bavaria | 1,458 | 15.84 | 742 | 15.85 |
| state | Saarland | 120 | 1.26 | 76 | 1.56 |
| state | Berlin | 333 | 3.39 | 169 | 3.50 |
| state | Brandenburg | 718 | 2.69 | 341 | 2.37 |
| state | Mecklenburg-West Pomerania | 438 | 1.82 | 188 | 1.46 |
| state | Saxony | 1,005 | 4.11 | 539 | 4.38 |
| state | Saxony-Anhalt | 582 | 2.47 | 289 | 2.38 |
| state | Thuringia | 608 | 2.31 | 328 | 2.49 |

Please note, the total sample refers to the KiGGS population with laboratory results. NA = no answer. Color code: green - difference less than 1 percent, orange - 1 to 2 percent, red - 2 to 3 percent between the total and final sample.

**Supplementary Table 6 B**

Comparison Between Total and Final Sample Concerning Important Characteristics Determining Representativeness Stratified by Sex

| variable | sex | label | $N_{total\ sample}$ = 11.024 | Proportion (%) | $N_{Final\ sample}$ = 5.559 | Proportion (%) |
|---|---|---|---|---|---|---|
| age group | male | 3-6 years | 449 | 7.64 | 208 | 6.71 |
| age group | male | 7-10 years | 1,929 | 30.70 | 964 | 28.39 |
| age group | male | 11-13 years | 1,487 | 24.42 | 913 | 27.22 |
| age group | male | 14-17 years | 1,801 | 37.25 | 965 | 37.68 |
| age group | female | 3-6 years | 412 | 7.65 | 194 | 7.72 |
| age group | female | 7-10 years | 1,810 | 30.70 | 830 | 30.54 |
| age group | female | 11-13 years | 1,407 | 24.46 | 706 | 25.11 |
| age group | female | 14-17 years | 1,729 | 37.20 | 779 | 36.63 |
| educational level | male | basic education | 972 | 34.11 | 507 | 33.33 |
| educational level | male | intermediate education | 3,041 | 43.57 | 1,668 | 44.68 |
| educational level | male | higher education | 1,541 | 20.47 | 821 | 20.39 |
| educational level | male | NA | 112 | 1.85 | 54 | 1.61 |
| educational level | female | basic education | 908 | 34.19 | 416 | 33.63 |
| educational level | female | intermediate education | 2,933 | 44.96 | 1,398 | 46.33 |
| educational level | female | higher education | 1,418 | 19.43 | 651 | 18.73 |
| educational level | female | NA | 99 | 1.41 | 44 | 1.32 |
| state | male | Schleswig-Holstein | 169 | 3.56 | 102 | 3.93 |
| state | male | Hamburg | 69 | 1.75 | 46 | 2.25 |
| state | male | Lower Saxony | 423 | 10.52 | 260 | 12.02 |
| state | male | Bremen | 33 | 0.73 | 17 | 0.72 |
| state | male | Northrhine-Westphalia | 1,091 | 23.11 | 609 | 24.21 |
| state | male | Hesse | 277 | 7.37 | 173 | 8.07 |
| state | male | Rhineland Palatinate | 242 | 5.21 | 88 | 3.70 |
| state | male | Baden-Württemberg | 677 | 13.96 | 300 | 10.99 |
| state | male | Bavaria | 751 | 15.84 | 406 | 15.95 |
| state | male | Saarland | 75 | 1.26 | 54 | 1.68 |
| state | male | Berlin | 163 | 3.29 | 90 | 3.53 |

| | | | | | | |
|---|---|---|---|---|---|---|
| state | male | Brandenburg | 361 | 2.70 | 175 | 2.29 |
| state | male | Mecklenburg-West Pomerania | 238 | 1.84 | 104 | 1.34 |
| state | male | Saxony | 501 | 4.08 | 293 | 4.50 |
| state | male | Saxony-Anhalt | 279 | 2.49 | 151 | 2.40 |
| state | male | Thuringia | 317 | 2.30 | 182 | 2.39 |
| state | female | Schleswig-Holstein | 155 | 3.56 | 89 | 4.68 |
| state | female | Hamburg | 67 | 1.76 | 32 | 1.69 |
| state | female | Lower Saxony | 437 | 10.60 | 236 | 12.19 |
| state | female | Bremen | 31 | 0.68 | 8 | 0.39 |
| state | female | Northrhine-Westphalia | 988 | 23.16 | 488 | 24.84 |
| state | female | Hesse | 290 | 7.35 | 158 | 8.19 |
| state | female | Rhineland Palatinate | 205 | 5.07 | 77 | 4.01 |
| state | female | Baden-Württemberg | 608 | 13.84 | 204 | 10.18 |
| state | female | Bavaria | 707 | 15.84 | 336 | 15.71 |
| state | female | Saarland | 45 | 1.25 | 22 | 1.40 |
| state | female | Berlin | 170 | 3.49 | 79 | 3.46 |
| state | female | Brandenburg | 357 | 2.68 | 166 | 2.47 |
| state | female | Mecklenburg-West Pomerania | 200 | 1.80 | 84 | 1.60 |
| state | female | Saxony | 504 | 4.15 | 246 | 4.23 |
| state | female | Saxony-Anhalt | 303 | 2.44 | 138 | 2.34 |
| state | female | Thuringia | 291 | 2.31 | 146 | 2.61 |

Please note, the total sample refers to the KiGGS population with laboratory results. NA = no answer. Color code: green - difference less than 1 percent, orange - 1 to 2 percent, red - 2 to 3 percent between the total and final sample.
